# Preference heterogeneity for HIV pre-exposure prophylaxis care among gay, bisexual, and other men who have sex with men in the United States: a large discrete choice experiment

**DOI:** 10.1101/2024.05.30.24308102

**Authors:** Viraj V. Patel, Eli Andrade, Rebecca Zimba, Chloe Mirzayi, Chenshu Zhang, Michael Kharfen, Zoe Edelstein, Anthony Freeman, Rupali Doshi, Denis Nash, Christian Grov

## Abstract

**Background:** PrEP uptake among Black and Latino gay, bisexual, and other men who have sex with men (GBM) remains low in the United States. The design and implementation of PrEP delivery programs that incorporate the preferences of Black and Latino GBM may overcome barriers to uptake. We aimed to identify preferences for PrEP care among high-priority GBM in the U.S. with a large discrete choice experiment.

**Methods:** We conducted two discreet choice experiments (DCE) to elicit care preferences for (1) Starting PrEP and (2) Continuing PrEP care among GBM clinically indicated for PrEP. The DCE web-based survey was nested in a longitudinal cohort study of GBM in the U.S., implemented with video and audio directions among 16-49 year-old participants, not using PrEP, and verified to be HIV-negative. All participants were presented with 16 choice sets, with choices determined by BLGBM and PrEP implementation stakeholders. We calculated overall utility scores and relative importance and used latent class analyses (LCA) to identify classes within the Starting and Continuing PrEP DCE. Multivariable analysis was performed to identify factors associated with class membership.

**Findings:** Among 1514 participants, mean age was 32 years; 46·5% identified as Latino, 21·4% Black, and 25·2 White; 37·5% had an income less than USD $20,000. Two latent classes were identified for Starting PrEP: *Class 1* (n=431 [28·5%]) was driven by preference for more traditional in-person care, and *Class 2* (n=1083 [71·5%]) preferred flexible care options and on-demand PrEP. In a multivariable model, having a sexual health doctor (adjusted OR 0·7, CI 0·5, 0·9), having a primary care provider (OR 0·7,CI 0·5, 0·9, p= 0·023), and concerns over PrEP side effects (OR 1·1, CI 1·0,1·2, p= 0·003) were all associated with class membership.

**Interpretation:** The different preferences identified for PrEP care indicate the need for diverse care and formulation choices to improve PrEP uptake and persistence. Addressing these preferences and understanding the factors that shape them can inform the implementation of programs that increase PrEP uptake.

## Introduction

New HIV infections in the United States continue to be concentrated among key populations, with gay, bisexual, and other men who reported male-to-male sexual contact (GBM) comprising majority of all new infections.^1^ HIV prevention with pre-exposure prophylaxis (PrEP) with oral tenofovir-emtricitabine and now long-acting injectable cabotegravir is highly efficacious, increasingly widely available across the United States (US), and a key component of the global and U.S. ending the epidemic initiatives. Despite the potential of this biomedical HIV prevention tool, there are enduring structural barriers to a robust and sustained implementation of PrEP, particularly among structurally marginalized GBM such as Black and Latino GBM (BLGBM) at high priority for HIV prevention.^2–5^ Although intention to use PrEP has been high among BLGBM in the United States, significant gaps between intention and use persist^6–9^ and uptake remains suboptimal.^8^ Significant disparities persist in both uptake and continued use of PrEP across racial/ethnic groups in the US.^10,11^ These data suggest that traditional approaches to implement PrEP are inequitable and likely recapitulating barriers to uptake and continued use seen with HIV treatment, thus exacerbating disparities in new HIV infections incidence by race/ethnicity, socioeconomic status, age, and geography in the U.S.^12,13^

New strategies to improve uptake and persistence of PrEP among GBM at high priority for HIV prevention are needed. Numerous factors influence PrEP use among BLGBM in the United States,^5,14–20^ including care delivery strategies such as access to convenient and streamlined clinical care, affirming providers, PrEP formulation (e.g., oral daily, on-demand, or long-acting injectable), and cost – many of which could be addressed through improved design and implementation of PrEP delivery programs. Implementation of care strategies that prioritize preferences of diverse BLGBM groups may help to promote more equitable access and reduce disparities in PrEP use. However, there is a paucity of data about which specific strategies for PrEP care may be most preferred and important to engage diverse GBM not using PrEP in the U.S.. Robustly understanding such care preferences can guide programs to allocate resources to care strategies that may have the greatest potential impact.

One approach to robustly identify preferences for PrEP care is the use of Discrete Choice Experiments (DCEs).^21^ DCEs is a quantitative approach that can be used to systematically elicit product or program preferences in a priority population for healthcare,^22^ and are increasingly applied in HIV research.^23,24^ DCEs can facilitate the identification of product or program attributes important to the population of interest,^24–28^ and allow implementers to optimize real-world effectiveness by better matching the product or program attributes to those preferred by the priority population or the relative trade-offs individuals may be willing to make.

While DCEs are increasingly being used in HIV research, few studies have used DCEs or other methods to robustly identify and prioritize preferences and trade-offs for PrEP care delivery among diverse GBM groups in the U.S. In prior DCEs about PrEP program features among GBM in the U.S (N = 554),^29^ the sample was primarily white (>80%) – limiting its ability to identify potential differences in care preferences among diverse BLGBM groups, and was conducted in 2015 when PrEP had not yet become readily available across the U.S. The objective of the current study was to identify preferences for PrEP care initiation and continuation among diverse GBM groups at priority for HIV prevention across the U.S. but not using PrEP.

## Methods

To identify the most preferred PrEP care options among individuals not currently using PrEP and high priority for HIV prevention, we conducted a DCE among a sample of participants from the *Together 5,000* cohort study (T5K), an internet-based longitudinal study of a geographically diverse U.S. national sample of HIV-negative men, trans men, and trans women who have sex with men,^30^ 50·7% of whom identify as a racial/ethnic minority. Eligibility criteria for the parent T5K study enrolled individuals with high vulnerability for HIV acquisition, aged 16-49, and not using PrEP at the time of enrollment.^31^ T5K participants at baseline also had to test HIV negative, have ≥2 male sex partners in the past 3-months, and met one or more additional criteria: in the past 12 months, diagnosed with rectal STI or syphilis, shared injection drug use needles, or took HIV post-exposure prophylaxis; in the past 3 months, ≥1 receptive condomless anal sex (CAS), >2 insertive CAS, or methamphetamine use. T5K participants were recruited via ads on geosocial sexual networking apps and completed twice annual online surveys and annual HIV testing by self-sampling (Orasure HIV-1 specimen collection device, returned to lab by mail for analysis).

### Development and design of the DCEs

We selected attributes and their levels based on a review of the literature about PrEP barriers and facilitators among GBM in the U.S. and eliciting input via individual interviews and focus group discussions with stakeholders. We conducted 15 individual interviews with PrEP program implementers at two health departments (New York City and Washington D.C.), community-based organizations, and clinical providers. We also conducted three focus groups with BLGBM at high priority for HIV prevention and not on PrEP (recruited from the parent T5K study). Interviews took place via telephone or web-based video meetings and participants received a $35 online gift card. This process resulted in a final list of attributes and the corresponding levels (see Table 1).

**Table 1.** Attributes and levels for each PrEP care discrete choice experiment.

| **Discrete Choice Experiment 1: Starting PrEP** |  |
| --- | --- |
| **Attribute** | **Level** |
| How to start the PrEP conversation | Visit a general primary care office/clinic<br>Visit a sexual health center or STI Clinic<br>By phone, a video call (e.g., FaceTime), or text message<br>Visit a pharmacy and speak with the pharmacist |
| Appointment availability | Monday through Friday, 8:30am-5pm<br>24 hours a day, 7 days a week |
| Lab testing for HIV/STI | You collect samples at home and mail them in<br>You go to a clinic or lab |
| When you start the PrEP medications | Same day as appointment<br>2 to 3 days after the appointment |
| **Discrete Choice Experiment 2: Continuing PrEP** |  |
| Where you see a provider for PrEP services | LGBTQ-focused health center<br>General primary care clinic or office<br>Video call (e.g. FaceTime) with a doctor<br>Sexual health center or STI clinic |
| How to get HIV and STI testing every 3 months | You go to a clinic or lab<br>You collect samples at home and mail them in<br>You test yourself at home, with results available right away |
| Form of PrEP | You take a pill every day<br>You take pills only when you have sex<br>You take an injection in the butt every 2 months, at a pharmacy or clinic |
| Follow-up appointments with a healthcare provider | Every 3 months<br>Every 6 months<br>Every 12 months |
| Cost of care and medication | \$0 (No costs or copays)<br>\$20 every 3 months<br>\$100 every 3 months |

**Table 2.** Participant characteristics by latent classes for Starting and Continuing PrEP Care discrete choice experiments.

| Characteristic |  | Starting PrEP |  |  | Continuing PrEP |  |  |
| --- | --- | --- | --- | --- | --- | --- | --- |
|  | Total N (%) | Class 1: *In-person*<br>(N= 431)<br>n (%) | Class 2: *Virtual*<br>(N = 1083)<br>n (%) | *P* | Class 1: *Pills*<br>(N= 357)<br>n (%) | Class 2: *No Cost/Injectable*<br>(N= 1157)<br>n (%) | *P* |
| **Age mean (SD)** | 29.7 (7.8) |  |  |  | 29.6 (7.8) | 29.7 (7.9) | 0.7109 |
| **Population Density mean (SD)** | 7651.15 (13844) | 8252.8 (15286.6) | 7424.3 (13225.8) | 0.2985 | 7065.2 (13499.1) | 7831.4 (13949.2) | 0.3669 |
| **Race/Ethnicity** |  |  |  | **0.0002** |  |  | 0.5968 |
| Black | 324 (21.4) | 102 (31.4) | 223 (68.6) |  | 82 (25.3) | 242 (74.7) |  |
| Latino | 704 (46.5) | 223 (31.6) | 483 (68.4) |  | 156 (22.2) | 548 (77.8) |  |
| Multiple/Other | 103 (6.8) | 30 (29.1) | 73 (70.9) |  | 23 (22.33) | 80 (77.7) |  |
| White | 383 (25.3) | 75 (19.6) | 308 (80.4) |  | 96 (25.1) | 287 (74.9) |  |
| **Gender identity** |  |  |  | **0.0189** |  |  | 0.1951 |
| Not cis-gender male | 40 (2.6) | 18 (45.0) | 22 (55.0) |  | 6 (15.0) | 34 (85.0) |  |
| Cis-gender male | 1474 (97.4) | 413 (28.02) | 1061 (71.9) |  | 351 (23.8) | 1123 (76.2) |  |
| **Education** |  |  |  | 0.0696 |  |  | 0.5195 |
| High school or less | 250 (16.5) | 83 (33.2) | 167 (66.8) |  | 55 (22.0) | 195 (12.9) |  |
| More than high school | 1264 (83.5) | 348 (27.5) | 916 (72.5) |  | 302 (23.9) | 962 (76.1) |  |
| **Annual Income** |  |  |  | **<.0001** |  |  | 0.3837 |
| Less than \$20,000 | 567 (37.5) | 188 (33.2) | 379 (66.8) | | 131 (23.1) | 436 (76.9) | |
| \$20,000 - \$49,999 | 655 (43.3) | 191 (29.2) | 464 (70.8) | | 146 (22.3) | 509 (77.7) | |
| \$50,000 - \$99,999 | 236 (15.6) | 41 (17.4) | 195 (82.6) | | 65 (27.5) | 171 (72.5) | |
| \$100,000 or more | 56 (3.7) | 11 (19.6) | 45 (80.4) | | 15 (26.8) | 41 (73.2) | |
| **Has health insurance** |  |  |  | 0.6093 |  |  | 0.3892 |
| No | 512 (33.8) | 150 (29.3) | 362 (70.7) |  | 114 (22.3) | 398 (77.7) |  |
| Yes | 1002 (66.2) | 281 (28.0) | 721 (71.9) |  | 243 (24.3) | 759 (75.8) |  |
| **Housing instability in past year** |  |  |  | 0.069 |  |  | 0.2378 |
| No | 1335 (89.5) | 367 (27.5) | 968 (72.5) |  | 320 (23.9) | 1015 (76.0) |  |
| Yes | 157 (10.5) | 54 (34.4) | 103 (65.6) |  | 31 (19.8) | 126 (80.3) |  |
| **PrEP – ever prescribed** |  |  |  | 0.323 |  |  | 0.1102 |
| No | 1346 (88.9) | 376 (27.9) | 970 (72.1) |  | 322 (23.9) | 1024 (76.1) |  |
| Yes | 168 (11.1) | 53 (31.5) | 115 (68.5) |  | 31 (18.5) | 137 (81.5) |  |
| **Has a place to go to for sexual healthcare** |  |  |  | **0.0004** |  |  | 0.8474 |
| No | 477 (31.5) | 107 (22.4) | 370 (77.6) |  | 111 (23.3) | 366 (76.7) |  |
| Yes | 1037 (68.5) | 324 (31.2) | 713 (68.8) |  | 246 (23.7) | 791 (76.3) |  |
| **Has personal doctor/healthcare provider** |  |  |  | 0.0674 |  |  | 0.9736 |
| No | 773 (51.1) | 204 (26.4) | 569 (73.6) |  | 182 (23.5) | 591 (76.5) |  |
| Yes | 741 (48.9) | 227 (30.6) | 514 (69.4) |  | 175 (23.6) | 566 (76.4) |  |
| **My doctor knows I have sex with men (N = 741)\*** |  |  |  | **0.0124** |  |  | 0.0562 |
| No | 244 (32.9) | 60 (24.6) | 184 (75.4) |  | 68 (27.9) | 176 (72.1) |  |
| Yes | 497 (67.0) | 167 (33.6) | 330 (66.4) |  | 107 (21.5) | 390 (78.5) |  |
| **Comfort talking about sex with a doctor\* (N=741)** |  |  |  | **<0.0001** |  |  | 0.5438 |
| Very uncomfortable | 90 (12.1) | 22 (24.4) | 68 (75.6) |  | 70 (9.4) | 20 (2.3) |  |
| Uncomfortable | 125 (16.9) | 25 (19.8) | 101 (80.2) |  | 91 (12.2) | 35 (4.7) |  |
| Neither uncomfortable or comfortable | 152 (20.5) | 43 (28.1) | 110 (71.9) |  | 117 (15.7) | 36 (4.8) |  |
| Comfortable | 191 (25.8) | 56 (29.3) | 135 (70.7) |  | 144 (19.4) | 47 (6.3) |  |
| Very comfortable | 183 (24.7) | 80 (43.5) | 104 (56.5) |  | 148 (19.9) | 36 (4.8) |  |
| **Perceived hassle to complete paperwork for free PrEP** |  |  |  | 0.0555 |  |  | 0.6700 |
| No | 901 (59.5) | 273 (30.3) | 628 (69.7) |  | 209 (23.2) | 692 (76.8) |  |
| Yes | 613 (40.5) | 158 (25.8) | 455 (74.2) |  | 148 (24.1) | 465 (75.9) |  |
| **Has used local health department for sexual healthcare** |  |  |  | **0.0008** |  |  | 0.0889 |
| No | 442 (29.2) | 99 (22.4) | 343 (77.6) |  | 117 (26.5) | 325 ( ) |  |
| Yes | 1072 (70.8) | 332 (30.9) | 740 (69.0) |  | 240 (22.4) | 832 (77.6) |  |
| **Anticipated Provider Stigma mean (SD)** | 2.41 (0.98) | 2.22 (0.98) | 2.48 (0.93) | **<0.0001** |  |  |  |

\*Question only asked to those who responded Yes to having a personal doctor

**Table 3.** Multivariable analysis of factors associated with preference class for starting and continuing PrEP.

|  | **Starting PrEP**<br>(reference: Latent Class 1: *In-Person care*) |  | **Continuing PrEP**<br>(reference: Class 1: *No cost/prefer pills*) |  |
| --- | --- | --- | --- | --- |
| **Variable** | **aOR (95% CI)** | **P** | **aOR (95% CI)** | **P** |
| Population Density | 1.0 (1.0, 1.0) | 0.9409 | 1.0 (1.0, 1.0) | 0.6230 |
| Age | 1.0 (0.9, 1.0) | 0.5864 | 1.0 (0.9, 1.0) | 0.2447 |
| Race/Ethnicity |  |  |  |  |
| Black (Non-Hispanic) | **0.6 (0.41–0.86)** | **0.005** | 0.8 (0.5, 1.5) | 0.4414 |
| Latino | **0.55 (0.4–0.75)** | **<0.001** | 1.0 (0.6, 1.7) | 0.4931 |
| Multiple/Other | 0.69 (0.41–1.16) | 0.16 |  |  |
| White | 1 (ref) |  | 1 (ref) |  |
| Cisgender-male (ref: not cisgender male) | 1.8 (0.9, 3.5) | 0.0731 | 0.6 (0.2, 1.5) | 0.2771 |
| Annual Income |  |  |  |  |
| <\$20k | **0.5 (0.2, 1.0)** | **0.0018** | 1.4 (0.7, 1.5) | 0.2738 |
| \$20,000–\$50,000 | 0.6 (0.3, 1.3) | 0.0615 | 1.4 (0.7, 2.7) | 0.1603 |
| \$50,000–\$100,000 | 1.1 (0.5, 2.4) | 0.7438 | 1.0 (0.5, 2.0) | 0.3351 |
| >\$100,000 | reference | | reference | |
| Education |  |  |  |  |
| High school or less | 0.8 (0.59, 1.1) | 0.169 | 1.0 (0.7, 1.5) | 0.6901 |
| More than high school | reference |  | reference |  |
| Ever used PrEP | 0.86 (0.65–1.15) | 0.309 | **1.4 (1.0, 2.0)** | **0.0153** |
| Has health insurance | 0.99 (0.76–1.3) | 0.961 | 0.9 (0.7, 1.3) | 0.8919 |
| Had Past year housing instability | **0.67 (0.47, 0.96)** | **0.028** | 1.1 (0.7, 1.8) | 0.4578 |
| No sexual health provider (ref: yes) | 1.26 (0.95–1.68) | 0.111 | 0.8 (0.6, 1.1) | 0.3606 |
| No primary care provider (ref: yes) | 1.28 (0.98–1.66) | 0.068 | 1.0 (0.7, 1.3) | 0.9791 |
| Visited health department for sexual health care (ref: did not visit) | 0.79 (0.6–1.04) | 0.086 | 1.1 (0.9, 1.5) | 0.2014 |
| Anticipated stigma from provider | **1.24 (1.09–1.42)** | **0.001** |  |  |
| PrEP barrier: concern speaking to doctors about PrEP | **1.1 (1.0, 1.2)** | **0.011** | **0.9 (0.8, 0.9)** | **0.0213** |
| PrEP barrier: stigma about PrEP use | 1.0 (0.9, 1.0) | 0.9787 | **1.0 (1.0, 1.1)** | **0.0129** |

We developed two different DCEs to focus on starting PrEP (four attributes) and continuing PrEP (five attributes), as programs may need to focus on different care attributes to support starting PrEP compared to continuing PrEP (i.e., ongoing or sustained use of PrEP after initiation). Additionally, having participants compare 9 attributes choices would have made the DCE overly complex and likely produced unreliable results. Each DCE had 16 choice tasks. See Figure 1 for an example choice task. Each choice task contained two juxtaposed scenarios comprised of different combinations of PrEP care features (i.e., attributes and levels) from which the participant had to select the preferred option. We randomized the combinations presented and the order of their presentation to each participant to reduce bias. The DCE was designed and implemented using Lighthouse Studio 9·8.1 (Sawtooth Software) and deployed using Sawtooth’s online survey hosting platform.

**Figure 1.**
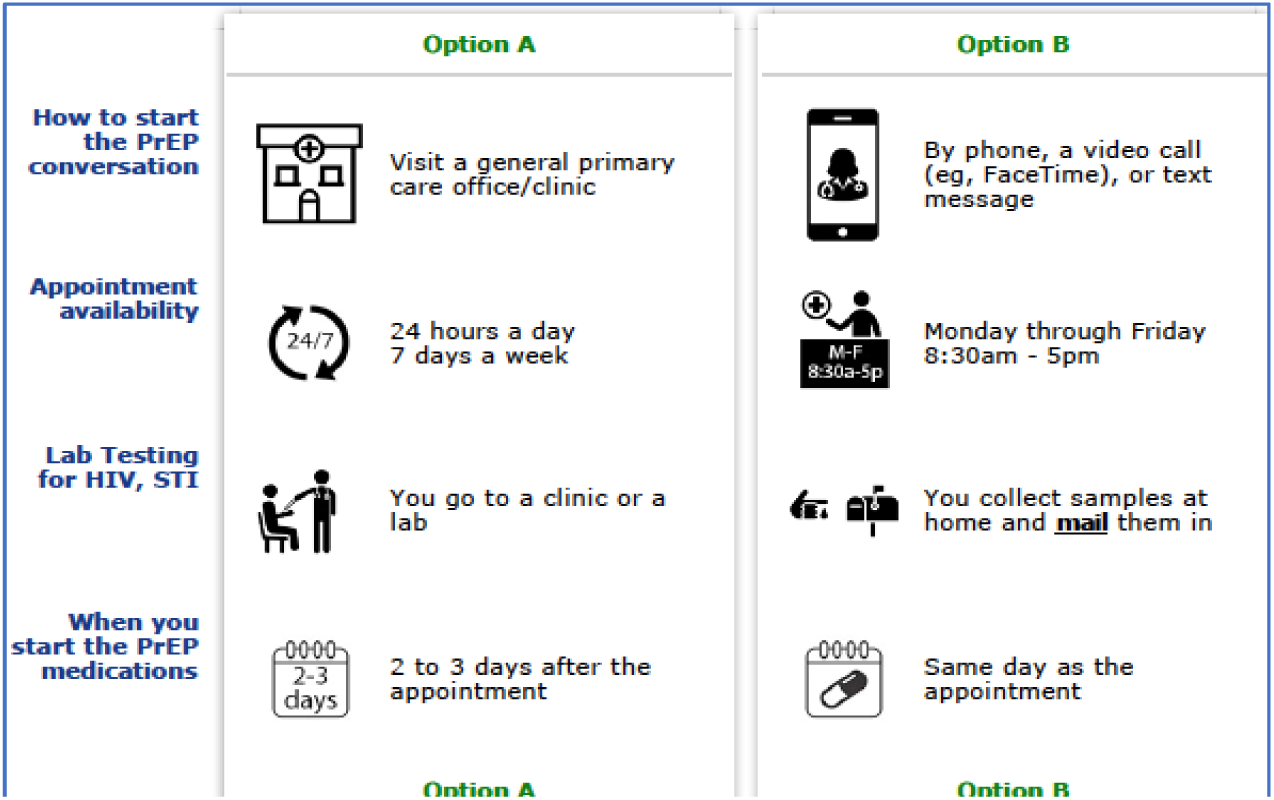
Example choice task for Starting PrEP.

### Study population and procedures

We aimed to enroll 1,500 T5K participants in the web-based DCE who, during their most recent follow-up assessment (either 12- or 24-month), tested HIV-negative and reported not using PrEP in the last 30 days. We stratified sampling by race/ethnicity (Black, Latino, and White) to help ensure robust comparisons between and within groups. Although the minimum sample size for DCEs is dependent on the maximum number of levels for any attribute, the number of choices in each choice set, and the number of choice tasks presented to each participant, precise power calculations can be difficult.^32^ As such, we determined our sample size to help ensure that robust comparisons would be possible between subgroups of interest, including by race/ethnicity, age, and history of PrEP use (i.e., PrEP-naïve vs prior use), as precise estimates for main effects in DCEs are generally optimized with sample sizes of about N = 200 per group.^33^

Invited participants received a personalized unique link via SMS or email to complete screening questions about their most recent HIV test and any recent PrEP use, as some individuals’ testing may have changed since their last T5K study assessment. Eligible participants who affirmed they were not HIV positive and had not used PrEP in the past 30 days PrEP at the time of the DCE then completed a web-based informed consent. Following consent, participants were provided with audiovisual and written directions for completing the DCE, a practice choice task about ice cream, one DCE about starting PrEP and a second about continuing PrEP, and a series of follow-up questions about health insurance, primary care, and stigma. We also merged information about participants’ demographic characteristics from the parent T5K study. Participants received a $25 online gift card for their participation.

### Latent Class Analysis

We estimated individual-level zero-centered part-worth utilities for each attribute level and overall relative attribute importance using a hierarchical Bayesian multinomial logit model (MNL). Next, we conducted a latent class analysis to characterize potential preference heterogeneity and identify which combinations of levels may be most important to implement for starting PrEP and then for continuing PrEP. For each DCE, two to five class solutions were explored. Model fit was assessed using common indices (Log-likelihood, Akaike’s information criterion [AIC], Bayesian information criterion [BIC]). We also considered clinical relevancy and interpretability of the results as a factor in our final solution. These analyses were performed using Lighthouse Studio 9·8·1 (Sawtooth Software).

#### Multivariable Regression analysis

Bivariate analyses using Chi-square and t-tests assessed associations between the latent class membership and predictors (e.g. socio-demographic characteristics, sexual behaviors). We then conducted a multivariable logistic regression analysis to estimate independent associations between participant characteristics and preferences (i.e. class membership) for Starting PrEP and Continuing PrEP. All factors used in the bivariate analyses were also included in the multivariable model. We tested for variance inflation factors (VIF) to examine multicollinearity among variables. A two-sided alpha level of 00·5 was used to determine significance for all analyses. The analysis was conducted in SAS (Version 9·4) and RStudio.

#### Ethical Review

This study was approved by the Institutional Review Board at the City University of New York (CUNY) and the Albert Einstein College of Medicine.

## Results

A total of 1512 individuals completed the DCE from March 2 - May 8, 2020, among whom the mean age was 29·7, most (97·4%) were cisgender men, 21·4% identified as Black (Non-Hispanic), 46·5% as Latino, and 25·2% White. The majority had college-level education or more (83·6%), 37% reported an annual income equal to or less than USD $20,000, and over a third did not have health insurance (33·8%). Over half (51·1%) did not have a personal doctor or healthcare provider and among those with a personal provider, about a third (244/741) reported their doctor or healthcare provider did not know they had sex with men.

### Starting PrEP Discrete Choice Experiment

The overall average importances of each attribute for Starting PrEP are shown in Figure 2a. The attribute “How to start the PrEP conversation” had the largest relative importance (43·0%), followed by “Where to get lab testing for HIV, STIs” (23%).

**Figure 2a.**
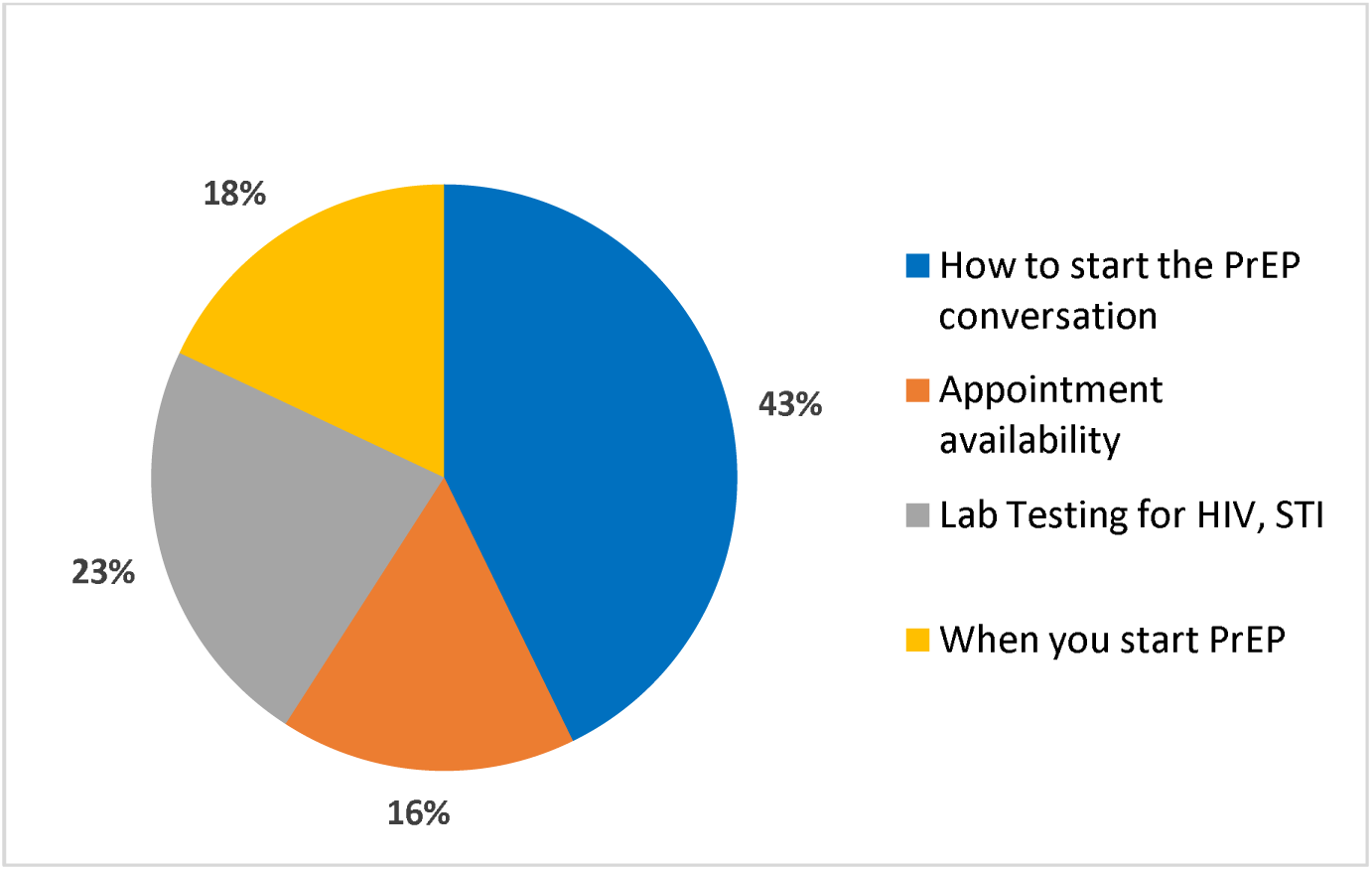
Starting PrEP average overall importances.

A two-class solution was identified for Starting PrEP. Class 1 (n = 431, 28·5%) was characterized by a traditional PrEP clinical care model (i.e., hereafter referred to as “In-person”). Class 2 (n = 1083, 71·5%) was characterized by preferring virtual or at-home care approaches (hereafter referred to as “Virtual”). The relative care preferences (i.e., zero-centered utilities) for each level of the Starting PrEP attributes are shown in Figure 2b by latent classes. The *In-Person* latent class preferred in-person visits for starting a PrEP conversation at a sexual health clinic followed by a general primary care clinic and for obtaining labs in-person. The *Virtual* latent class had a stronger preference for starting the the PrEP conversation with virtual care options and at-home self-sample collection for lab testing. Both groups preferred flexible appointment availability and same-day PrEP start.

**Figure 2b.**
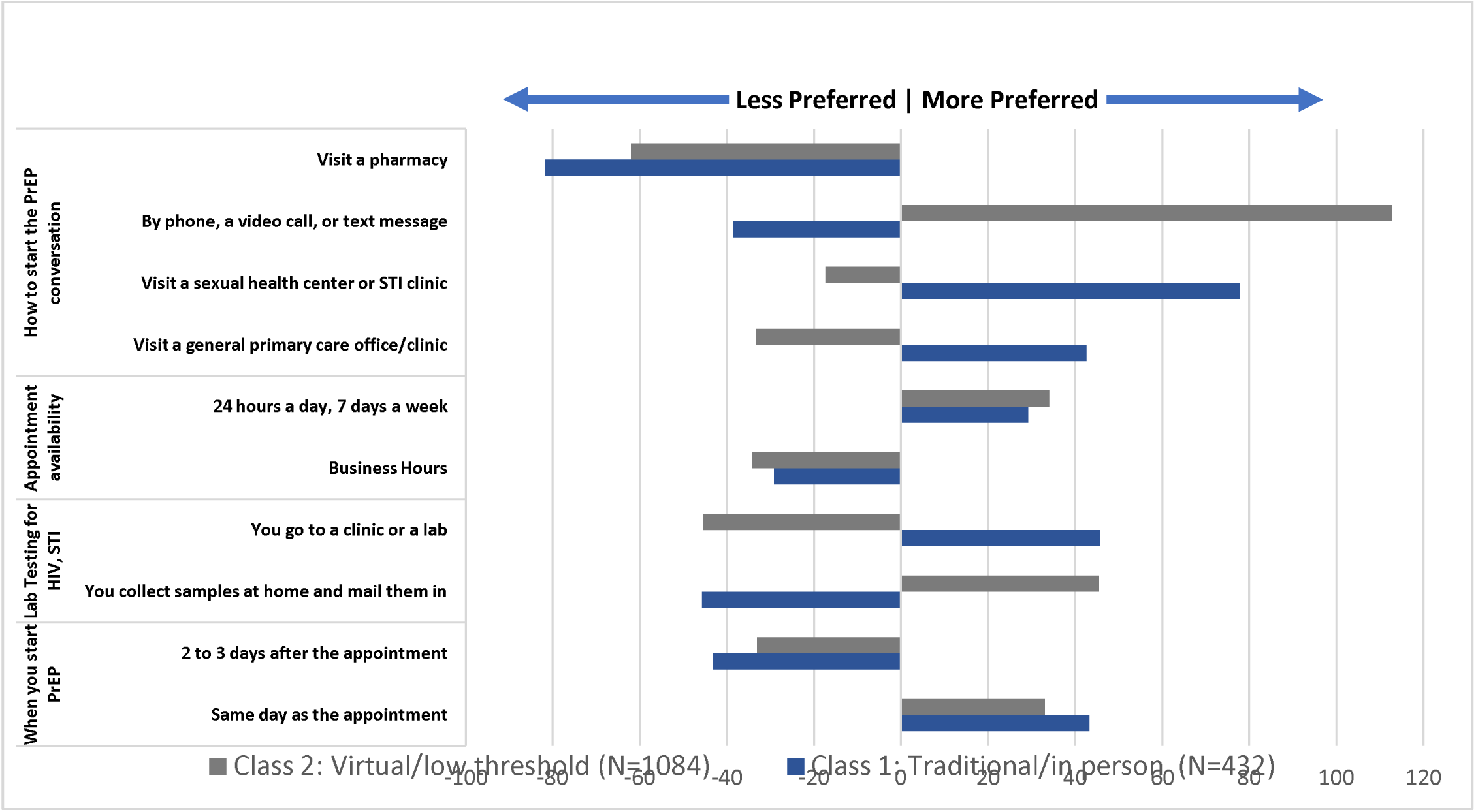
Relative Care Preferences for Starting PrEP by Latent Class.

### Continuing PrEP Discrete Choice Experiment

A two-class solution was also identified for Continuing PrEP. Class 1 was characterized by preferring low-cost options and PrEP in pill-form. Class 2 was characterized by preferring no-cost options and injectable PrEP. For continuing PrEP, the attributes “Cost of care and medication” and “Form of PrEP” had the largest importance levels, 26% and 58%, respectively (see Figure 3a). The most important attribute relative to all other choices was cost, and specifically a very strong preference for no cost or copays. The next most important driver of continuing PrEP was the formulation. Class 1 had a strong preference for oral pills using on-demand dosing and a strong negative preference for an every 2-months injectable PrEP. Group 2 had a preference for injectable PrEP and a strong negative preference for daily oral PrEP.

**Figure 3a.**
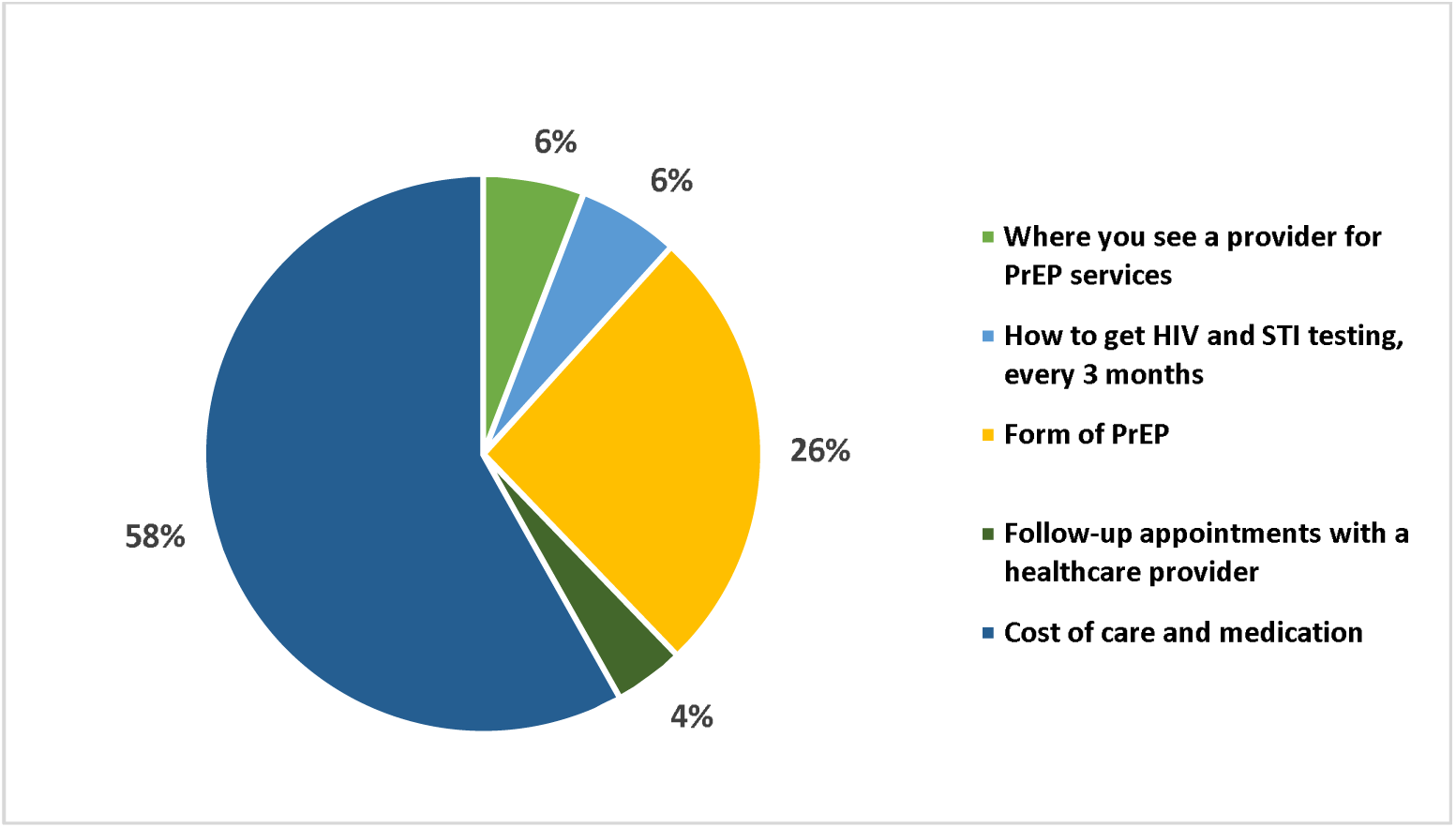
Continuing PrEP average overall importances.

**Figure 3b.**
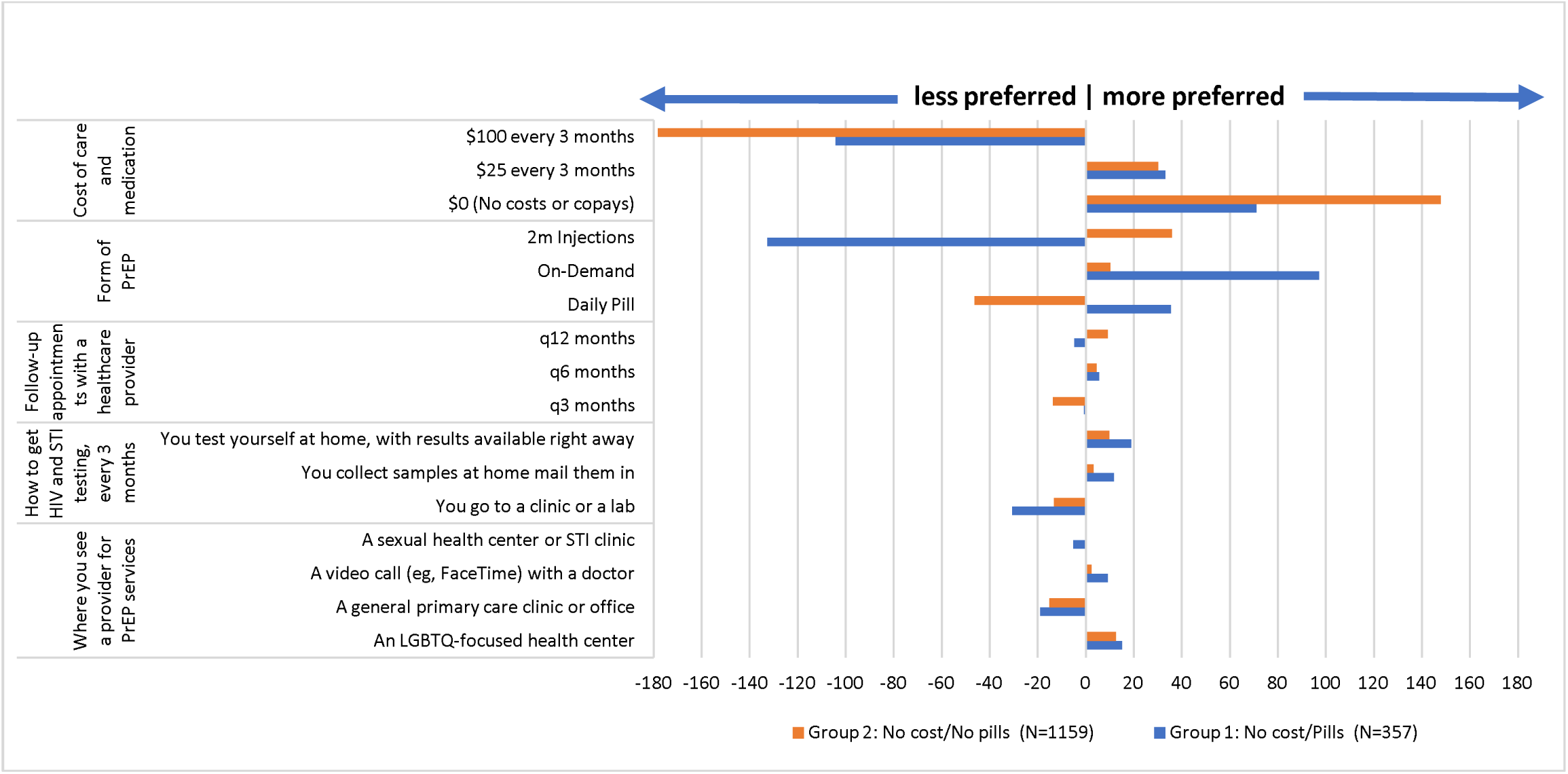
Care Preferences for Continuing PrEP by Latent Classes.

### Multivariable Analyses

In the Starting PrEP multivariable model, the virtual care group (Class 2) compared to the In-Person/Traditional care group (Class 1), had lower odds (AOR, 95% CI, all *P*<.05) of having access to a sexual health care (AOR 0·7, 0·5-0·9), having a primary care provider (AOR 0·7, 0·5-0·9), and higher odds of concerns about PrEP side effects (AOR 1·1, 1·0-1·2). For Continuing PrEP, Group 2 (reference: Group 1) had higher odds of having ever used PrEP (AOR 0·9, 1·0-2·0) and PrEP stigma as a barrier (AOR 1·0, 1·0-1·1).

## Discussion

In this discrete choice experiment study with a large geographically diverse sample of GBM from across the U.S. with behaviors at high risk for HIV, there was significant heterogeneity in preferences for care options for starting and continuing PrEP. In this study, we found that the strongest preference drivers were virtual versus in-person care options, having no cost for care visits or medications, and choice of PrEP formulation (on-demand and daily pills, long-acting injectable) and that these attributes drove stated preferences with relatively small contributions from other care options. Our findings underscore the need to provide choices for PrEP care, particularly beyond the traditional face-to-face clinical care models that predominate. For starting PrEP, the majority of participants in this sample had a strong preference for virtual care options (i.e., telemedicine and at-home labs) but almost one-third still preferred in-person care. Meanwhile for the Continuing PrEP DCE, no or low cost options and access to choices of PrEP formulations strongly drove preferences for PrEP use. Other aspects for PrEP care, such as lab testing location, time to starting PrEP, and frequency of follow-up appointments had diverse and diverging preferences, but were of relatively minor importance in this study. PrEP implementation programs could consider offering choices responsive to these preferences to optimize engaging and retaining diverse GBM at high priority for HIV prevention in PrEP care.

The results for the first DCE (Starting PrEP) showed a majority of the sample (71·5%) preferred virtual PrEP access, as well as at-home lab testing and same day PrEP start. Technology-based and low-threshold PrEP programs (i.e. telemedicine, virtual PrEP counselors, same day PrEP start) have been found to be acceptable, safe and feasible for PrEP uptake and retention in care among GBM.^34,35^ These strategies have been used to address barriers to PrEP uptake such as geographical location, time availability, need for in-person visits, reducing stigma, and addressing privacy concerns^36,37^ and expanding the use of such tools may improve uptake. However, we still found that a substantial proportion of GBM in this study (28·5%) preferred in-person care. These participants were more likely to identify as Black or Latino or have lower income. Embedding flexibility in PrEP delivery programs by leveraging technology, while also making access to in-person care easier, may address barriers at multiple levels by offering diverse paths for accessing PrEP care that meet the needs and preferences of diverse GBM in the U.S.. Additionally, this study was implemented at the time COVID-19 cases were rising, which may have also influenced preferences for virtual access.

Cost of PrEP and PrEP care continues to be a key barrier for accessing this effective prevention strategy in the United States. Our findings indicate that eliminating patient costs for PrEP care may have the largest impact on PrEP use relative to other factors examined. A recent review found that when cost sharing was eliminated for preventive care, uptake increased especially for those who had lower income.^38^ Cost barriers are likely compounded by the fragmented structure of the healthcare system, insurance coverage tied to employment and income-level, varied Medicaid expansion across states, and fragmented PrEP assistance programs (e.g., PrEP-AP in New York State which covers clinical and lab costs associated with PrEP care, and other programs cover medication costs).^39^ GBM in states with Medicaid expansion were more likely to have insurance, discuss PrEP with a provider, and to use PrEP.^40^ The majority of the states that have not expanded Medicaid are located in the south, where inequities in healthcare access and racial/ethnic disparities in HIV infections persist.^40^ For those who are insured, change or disruptions in insurance status has been associated with PrEP discontinuation due to inability to cover cost of the medication and associated care.^41,42^ While assistance programs such as PrEP-AP may cover the cost of provider and laboratory testing visits and medications, as of 2021, only 14 states had such programs with varying levels of cost coverage. Thus, policies and programs to reduce – if not eliminate – costs for PrEP are needed.

Our findings suggest that implementing on-demand and long-acting injectable PrEP may improve uptake among GBM who are not using PrEP. Heterogenous preferences for PrEP regimens among a large sample of GBM not using PrEP underscore the need to implement and promote access to options beyond a daily pill and may also reflect temporal fluctuations in the behaviors of GBM.^43^ On-demand or episodic PrEP (taking pills before and after sex) is highly effective for HIV prevention and non-inferior to daily pills, and has been shown to be acceptable for GBM who prefer not to take a daily medication.^44,45^ A recent study reported high levels of adherence among GBM using episodic-PrEP,^46^ a regimen option that can meet the preferences of GBM not interested in daily-PrEP. Additionally, long-acting injectable PrEP with cabotegravir has been found to be highly efficacious and even superior to oral formulations (possibly driven by adherence) among multiple populations (GBM, trans- and cis-gender women)^47,48^ as well as acceptable to GBM.^49–52^ Providing choices for formulation, especially combined with eliminating out of pocket costs to patients, may have a large impact on uptake and merit examination.^53^ Findings from contraception studies demonstrate that when cost and access issues were removed, women had high rates of contraception uptake and persistence (particularly with long acting options) and decreased unintended pregnancies,^54^ suggesting that a similar pattern of uptake and decreased HIV infections may hold true for PrEP implementation as well.

Interestingly, we observed in the *Starting PrEP* DCE that individuals who reported more anticipated stigma from a provider or had concerns about speaking to doctors about PrEP, had a higher preference for starting the PrEP conversation in more non-personal ways (e.g., telehealth) and at-home lab sampling and testing. Studies show that anticipated stigma represent barriers for PrEP uptake and can contribute to high-levels of non-disclosure of sexual-orientation, behaviors, and substance use to healthcare providers.^55,56^ Additionally, medical mistrust – encompassing mistrust of healthcare providers, health systems, and pharmaceutical medications – has been associated with disparities in HIV prevention and treatment use, particularly impacting populations at highest HIV risk^20,57,58^ and may be characterized in part by anticipated stigma and communication concerns.^59^ For example, among Black GBM, medical mistrust can be a barrier for routine preventive medical care,^60^ while for Latino GBM it has been associated with lower outcomes across the PrEP cascade.^20^ Our findings suggest that virtual care options may be a strategy to help overcome anticipated stigma related to sexual behaviors and identity, medical mistrust, reduce provider communication barriers and potentially improve PrEP uptake.^61,62^

## Limitations

Findings from this study should be interpreted in the context of its limitations. First, not all potential care attributes were able to be tested in this study and there may be other drivers not measured. However, we selected attributes based on a systematic review of PrEP barriers and facilitators^5^ and input from BLGBM meeting CDC eligibility for PrEP, planners from two health departments, and PrEP program implementers. Next, DCE are hypothetical, and it is possible that the stated preferences may not directly predict actual behavior; a systematic review and meta-analysis of DCEs found that reported preferences generally do correspond with actual behaviors, but research is warranted to understand whether this holds true for PrEP as well. Although we had a large geographically and racially/ethnically diverse sample at high priority for HIV prevention, findings nevertheless may not be generalizable to all GBM in the U.S. who may benefit from PrEP, and there may be other preference patterns not captured by this study. The study was implemented amidst the first major wave of the COVID-19 pandemic in the U.S. (March – May, 2020). Our findings regarding strong preferences for telehealth medicine could be a factor of many social distancing and stay-at-home recommendations in effect at that time. We note however, that the COVID pandemic has had sustained dramatic shifts in facets of daily life, including sustained increased availability of and demand for telemedicine. Thus, the fact that our data were gathered after the start of the COVID pandemic (as opposed to before) is perhaps more emblematic of what could be colloquially referred to as “our new reality.”

## Conclusions

Results from this study underscore the need for certain choices in PrEP care, and that no one model of care will likely meet the needs of most GBM including BLGBM. Findings have implications for the design and implementation of PrEP delivery programs, and can provide guidance to policymakers and program implementers on where to consider channeling resources to facilitate PrEP uptake among GBM at the highest priority for HIV prevention in the United States. Given that the type of care access, cost, and formulation were the strongest preference drivers for starting and continuing PrEP, implementation of state and federal policies are likely needed to ensure equitable care access and cost coverage across the United States.

## Data Availability

All data produced in the present work are contained in the manuscript.

